# Developing Evidence-Based Criteria for Oxycodone Prescribing in Emergency Departments: A Protocol for the OxyGuidED Study Using the RAND/UCLA Appropriateness Method

**DOI:** 10.64898/2026.02.01.26345327

**Authors:** Suhair M Althagafi, Samantha Keogh, James Hughes

**Author notes:** Corresponding author at: School of Nursing, Queensland University of Technology, Brisbane, Australia.

## Abstract

**Background:** Oxycodone is widely prescribed for managing acute pain in emergency departments (ED), but the appropriateness of this prescription is not fully established. Although concerns about opioid misuse and dependence drive efforts to reduce inappropriate prescribing, there is increasing recognition of the importance of adequate pain management. Therefore, it is essential to develop appropriate prescribing criteria that balance the risks and benefits of opioids, ensuring their benefits are maximised while minimising potential harm.

**Objective:** Following the recommended format for a research protocol paper, this protocol describes the process and methods used to develop evidence-based criteria for oxycodone prescribing in the ED, informed by scientific evidence and expert clinical judgment, using the RAND/UCLA Appropriateness Method.

**Method:** The process will be carried out in sequential stages. First, scope and key terms will be defined, and then a targeted literature review will be conducted to synthesise available evidence. Subsequently, based on this synthesis and the investigator team’s clinical insights, clinical scenarios will be developed in collaboration with field experts. A multidisciplinary panel comprising specialists in emergency medicine, emergency nurses, and pharmacists will evaluate these scenarios in two rounds. Each scenario will be rated on a 1-9 scale, where 1 indicates that harm outweighs benefit and 9 indicates that benefit outweighs harm. The median rating score will fall between 1 and 9, where 1-3 without disagreement is inappropriate, 4-6 without disagreement is uncertain, and 7-9 without disagreement is appropriate. Disagreement is defined as at least three experts scoring in both extremes. Final scenario ratings will be presented according to their assessed appropriateness and used to inform appropriateness criteria for prescribing oxycodone in the adult ED.

**Conclussion:** The RAND/UCLA Appropriateness Method offers a systematic and evidence-informed framework for developing prescribing criteria to support the appropriate use of oxycodone in adult ED.

## 1. Introduction

Pain is a common complaint in the emergency department (ED), where opioids are frequently prescribed for their effective and rapid pain relief (Gleber et al., 2020; Pouryahya et al., 2020). In Australian EDs, oxycodone accounts for 23.9% of all prescribed analgesics, making it the most prescribed oral opioid (*Australian Institute of Health and Welfare*, 2019; Hughes et al., 2024; Nguyen et al., 2021; Pouryahya et al., 2020). It is effective and has fewer side effects than morphine, yet it poses a higher risk of abuse, as evidenced by higher likability scores compared to morphine and hydrocodone (Kibaly et al., 2021). Its accessibility and affordability make it appealing for prescribing, and increase the risk of potential drug diversion (Cicero et al., 2013; Kibaly et al., 2021). This underscores the need to scrutinise prescribing practices in all settings, including the ED.

The prescription of oxycodone in the ED lacks standardised protocols, as evidenced by the varying prescription rates across different patient groups and ED settings (Chabon et al., 2024; Mitra et al., 2023). Moreover, the strategy of restriction is effective in reducing the harm of specific treatments (Beckwith et al., 2002; Mitra and Cameron, 2006; Mitra et al., 2023). However, the long-term impact of restricted access or reduced oxycodone prescriptions on quality pain management remains unassessed, despite restrictions reducing short-term opioid use. Moreover, oxycodone remains a cheap, effective analgesic agent for acute pain and will remain a mainstay of pain care in the ED. However, clear guidance on which patients’ oxycodone may be appropriate for is also required to reduce the inappropriate prescription of this potentially dangerous medication.

Clinical decision-making in the ED is frequently characterised by time pressure, diagnostic uncertainty, and heterogeneous patient presentations, conditions under which guideline-based recommendations may be difficult to apply consistently. While clinical practice guidelines synthesise evidence at a population level, they often lack the granularity required to support case-based prescribing decisions. This limitation is particularly evident in high-variability opioid prescribing in acute care settings (Hoppe et al., 2017). This is coupled with the need for clinicians to rapidly balance analgesic effectiveness against patient-specific risk factors and potential harm in the absence of clear, context-sensitive criteria. Therefore, the criteria are needed to help determine the appropriateness of oxycodone prescription, accounting for the diverse population of the ED, and in consideration of both short- and long-term benefits while minimising harm. This approach may help prevent both inadequate pain management and inappropriate oxycodone prescription among ED patients.

The RAND/UCLA Appropriateness Method was selected to provide a structured, transparent approach for integrating the best available evidence with expert clinical judgment in areas where empirical data alone are insufficient to guide practice (Fitch et al., 2001). Applying the RAND/UCLA Appropriateness Method in the ED context enables the development of OxyGuidED prescribing criteria that reflect real-world clinical complexity while maintaining methodological rigour, reproducibility, and clarity for future evaluation and implementation.

This paper will outline each stage of the RAND/UCLA Appropriateness Method process used to develop the OxyGuidED criteria, based on Fitch et al. (2001) and following the recommended medical research protocol format (Taylor, 2018).

## 2. Method

To develop criteria for appropriate oxycodone prescription in adult patients presenting to the ED, this study employs the RAND/UCLA Appropriateness Method (Fitch et al., 2001). This validated method combines the best available scientific evidence with clinical expertise to assess the appropriateness of healthcare decisions. The criteria produced are context-specific, informed by both evidence and experience, and designed to support more consistent and appropriate prescribing practices. The phases and steps of the RAND/UCLA Appropriateness Method process are summarised in Figure 1, with detailed descriptions provided in the sections that follow.

**Figure 1:**
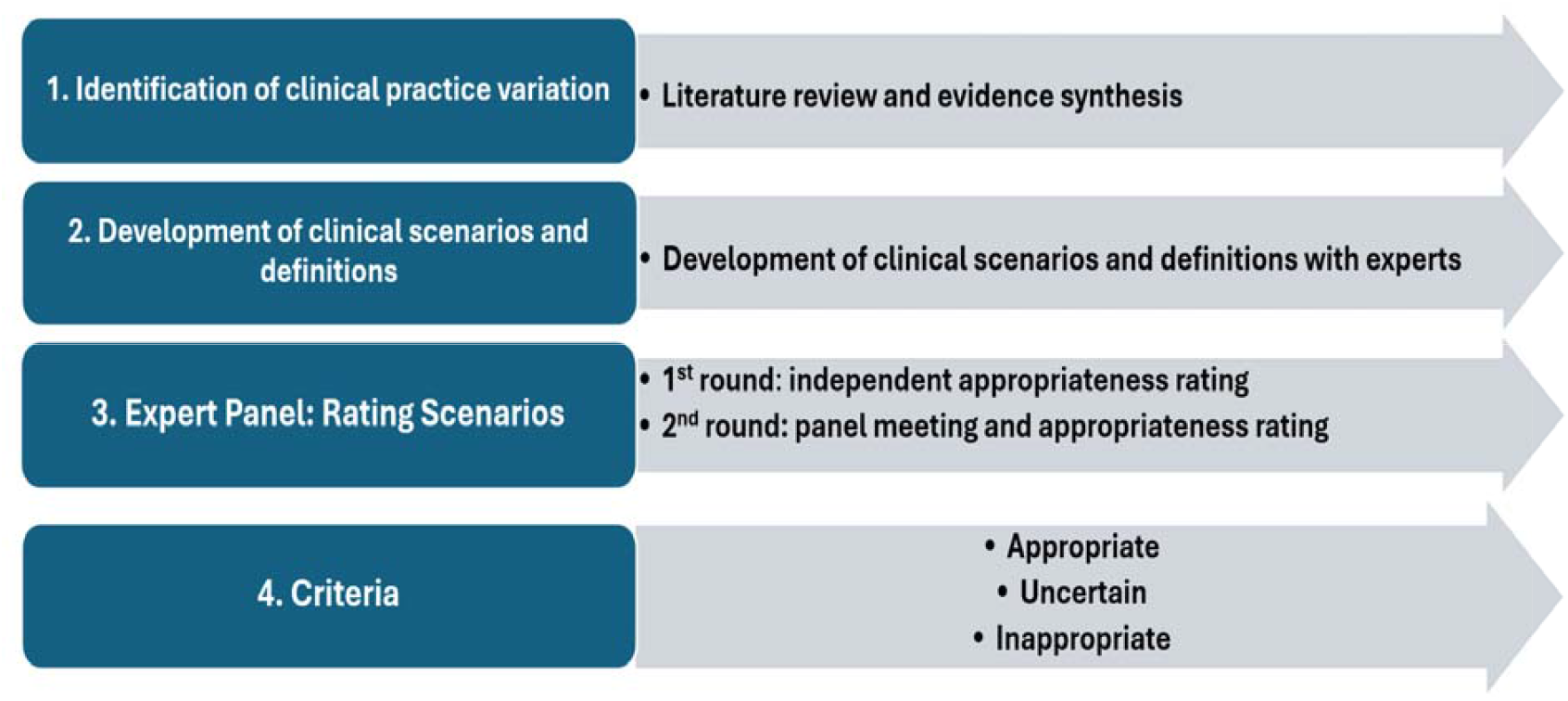
RAND/UCLA Appropriateness Method Phases

### 2.1. Evidence review and synthesis

The RAND/UCLA Appropriateness Method review is regarded as the foundation of the pyramid, which helps develop valid scenarios and aligns them more closely with real-world cases. Hence, an integrative review will be conducted to synthesise evidence related to optimising oxycodone use in adult ED patients. This review type is well-suited to incorporate multiple study designs and provide a comprehensive understanding, even when high-quality evidence is limited (Whittemore and Knafl, 2005). The review outcomes will inform the development of the scope, key terms, and clinical scenarios for the upcoming expert panel assessment.

### 2.2. Definition of scope and key domains

This stage in the RAND/UCLA Appropriateness Method focuses on clearly defining the scope of the clinical topic and pinpointing related key terms (Fitch et al., 2001). As previously discussed, oxycodone is commonly used in the ED for managing moderate to severe pain, but its overuse and some uncertainty about oxycodone prescribing carry many risks and harms. Therefore, the primary focus of the OxyGuidED study is to evaluate the clinical appropriateness of prescribing oxycodone for acute pain management in the adult ED.

This stage not only identifies the study’s scope but also organises the survey into main domains and further divides them into detailed chapters. The domains of the RAND/UCLA Appropriateness Method survey are broad, encompassing thematic areas identified from the literature or experience that are relevant to the intervention or decision being evaluated (Fitch et al., 2001). Recognising these domains will help shape the overall structure of the RAND/UCLA Appropriateness Method survey.

The major themes in the OxyGuidED survey will consider both clinical context and scientific evidence. Since the scope is to identify the boundaries of appropriateness of oxycodone within the ED, and considering that oxycodone is prescribed for moderate to severe pain, major domains in this survey will be related to both the context of oxycodone prescription within the ED and the indication for oxycodone prescription. These domains highlight two main aspects: the timing of oxycodone prescription (either during the ED visit or at discharge) and the reason for the ED visit for pain management (whether trauma-related, a new health issue, or ongoing health problems) (Figure 2). This classification system for the OxyGuidED survey will facilitate a comprehensive analysis of oxycodone use patterns across various clinical conditions.

**Figure 2:**
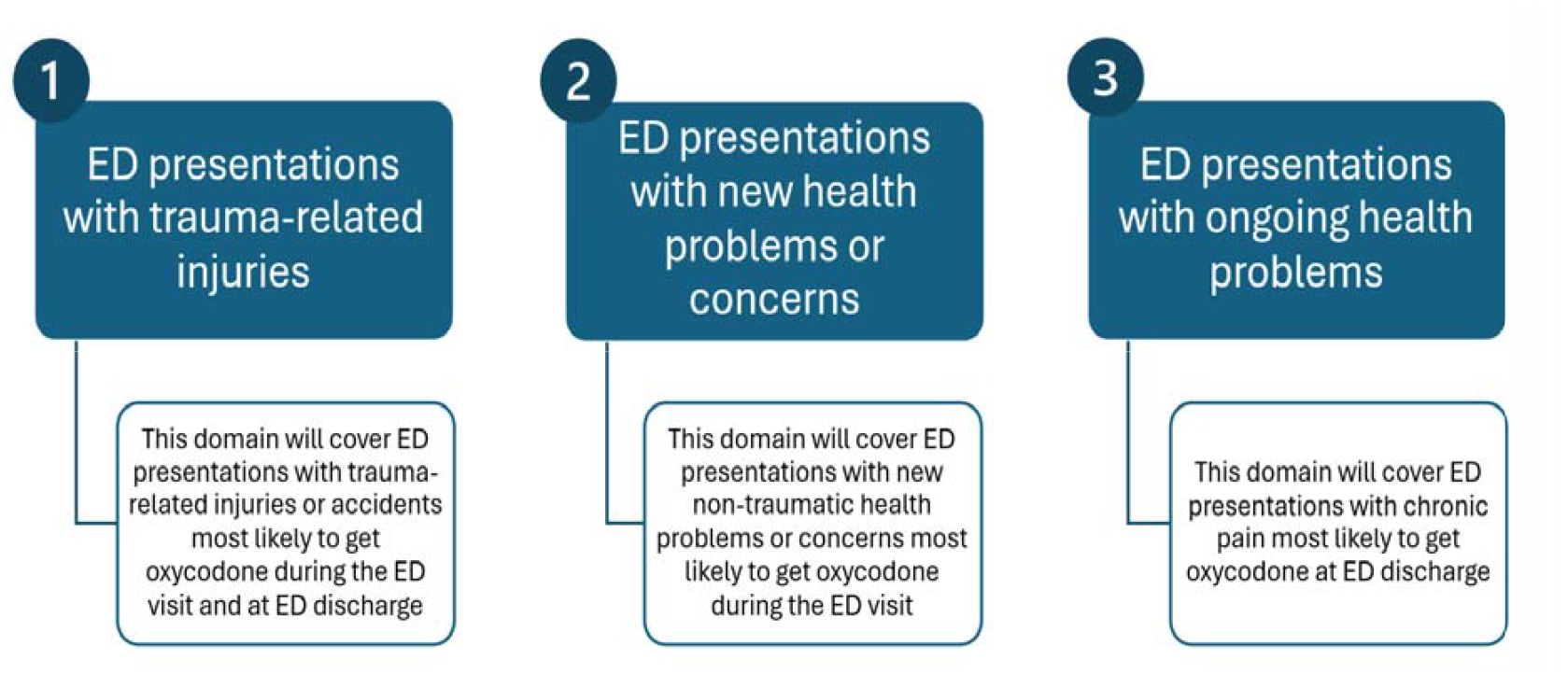
Major domains in the OxyGuidED survey

### 2.3. Categorisation of scenarios into chapters

Within each domain, previously listed, there are further divisions into chapters, which provide a more detailed view of the treatment or procedure of interest (Fitch et al., 2001). These chapters are designed to group scenarios by relevance, assisting panellists in rating, discussing, and interpreting the results.

Chapters in the OxyGuidED survey will be organised by body regions derived from The Barell Injury Diagnosis Matrix, Classification by Body Region and Nature of the Injury (CDC, 2005). Categorising the RAND/UCLA Appropriateness Method survey by body parts helps panellists make informed judgments about the suitability of oxycodone. This decision depends on the injured or painful areas, potential causes of pain, and other affected body systems (Figure 3). This detailed categorisation allows healthcare professionals and researchers to identify trends, compare practices, and assess the appropriateness of oxycodone prescriptions across various medical conditions, as well as anatomical areas.

**Figure 3:**
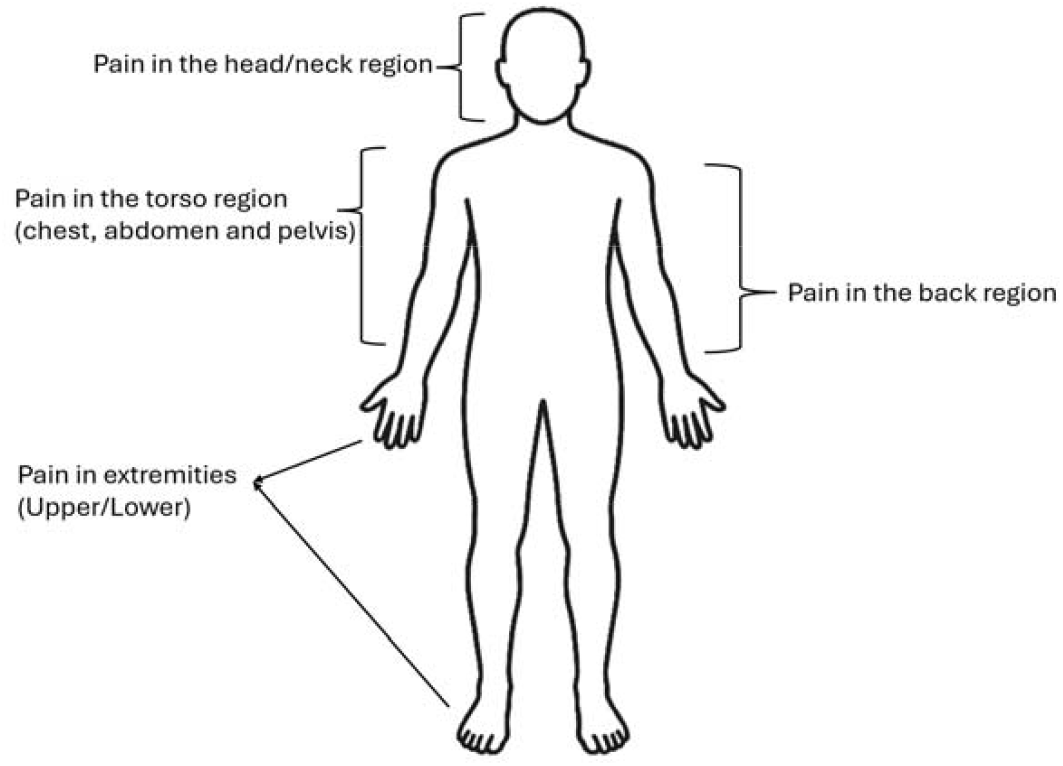
Categorisation of chapters by pain area in the OxyGuidED survey

### 2.4. Development of clinical scenarios

The RAND/UCLA Appropriateness Method survey will be developed through a streamlined process involving two rounds of clinical scenario assessments by panellists. It will identify uncertainties from current practices and the clinical literature, while incorporating expert insights. The scenario will be structured into chapters by relevance and focus to achieve a comprehensive approach.

The OxyGuidED study researchers aim to develop more targeted scenarios, limited to fewer than 300, with the rating process expected to take less than two hours. Within each chapter, a series of scenarios will be created, typically in the form of structured statements (see Table 1). These scenarios encompass multiple factors related to the treatment or procedure, capturing the complexity of real-world situations (Fitch et al., 2001). The list of key elements can be seen as factors that might influence the appropriateness of oxycodone prescriptions, such as patient age, patient condition, presence of chronic disease, and other factors reported in Figure 5. A clear definition of the highlighted key terms should be provided to the panellists along with the scenario to ensure a common understanding of the related terminology among panellists.

**Table 1:**
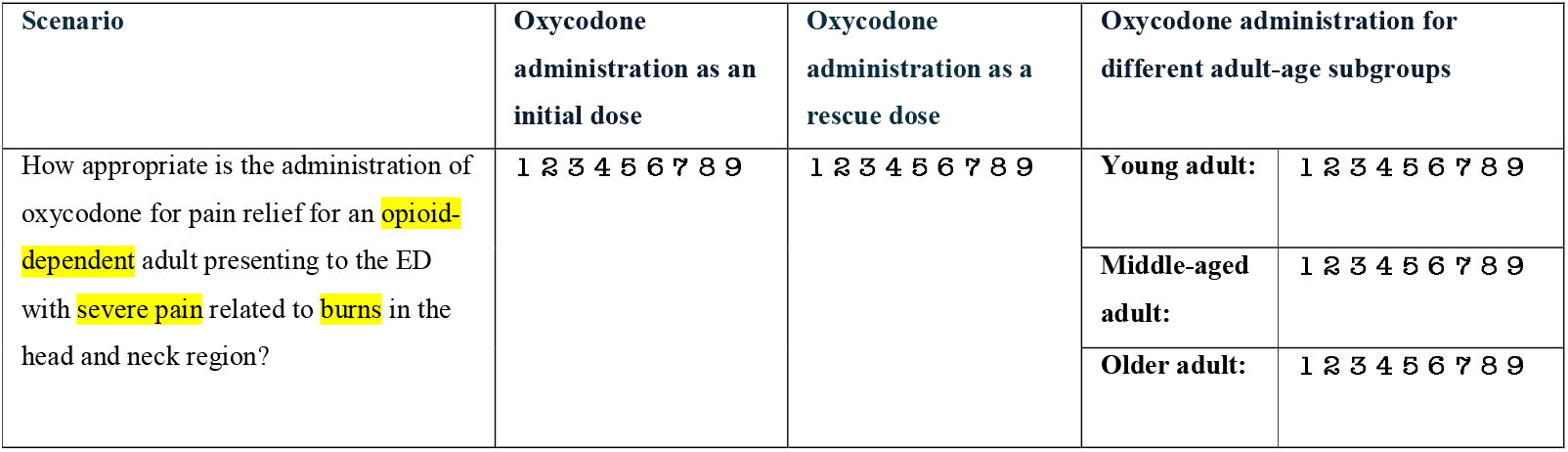
An example of a statement scenario in the OxyGuidED survey.

| Scenario | Oxycodone administration as an initial dose | Oxycodone administration as a rescue dose | Oxycodone administration for different adult-age subgroups |  |
| --- | --- | --- | --- | --- |
| How appropriate is the administration of oxycodone for pain relief for an opioid-dependent adult presenting to the ED with severe pain related to burns in the head and neck region? | 1 2 3 4 5 6 7 8 9 | 1 2 3 4 5 6 7 8 9 | Young adult: | 1 2 3 4 5 6 7 8 9 |
|  |  |  | Middle-aged adult: | 1 2 3 4 5 6 7 8 9 |
|  |  |  | Older adult: | 1 2 3 4 5 6 7 8 9 |

**Figure 5:**
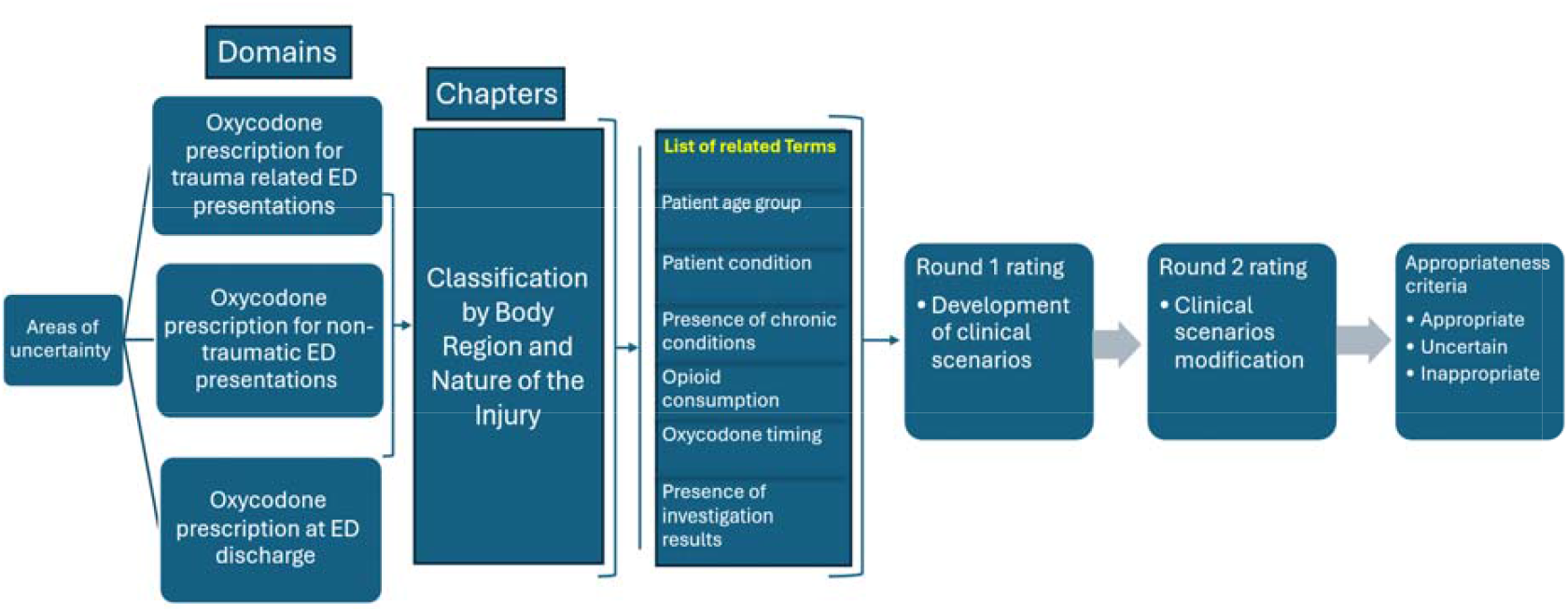
Conceptual framework for the development of clinical scenarios regarding the appropriateness of oxycodone use in the adult ED.

The inclusion of these related elements serves several purposes. First, it enhances the validity and relevance of the scenarios, thereby making them more applicable to real-world clinical decision-making. Secondly, discussing these scenarios promotes a more comprehensive assessment, encourages analytical thinking, and supports knowledge exchange among healthcare professionals. Additionally, the complexity of scenarios can be utilised for training, helping healthcare providers prepare for a wide range of clinical situations they may encounter. By reflecting the complexity of real-world settings, these scenarios enhance the quality of clinical decisions and, ultimately, patient care.

### 2.5. Refinement and validation process

The clinical scenarios to be developed will undergo an internal review process by both investigators and clinical experts to ensure alignment with the study’s predefined objectives (Fitch et al., 2001). Pilot testing will be conducted by experts who will not participate in the panel or scenario rating to provide clarity and relevance. This will include content validation of the clinical accuracy and completeness of scenarios across all possible oxycodone indications, as well as comprehension and wording relevant to real-world clinical terms and cases. Pilot testing will also include procedural validity to assess the feasibility of the rating process, as well as checks on data quality and scoring accuracy. The objective is to confirm the face validity of the RAND/UCLA Appropriateness Method survey, including consistent interpretation, standardised wording, and a stable data-collection process.

The collaborative core of the RAND/UCLA Appropriateness Method, which integrates scientific evidence and clinical expertise, serves several purposes. Firstly, it validates the content and structure of the scenarios, ensuring they accurately represent the clinical situations they aim to simulate. Secondly, it enables the refinement and enhancement of scenarios based on diverse professional insights, potentially uncovering nuances or considerations that may have been initially overlooked. Ultimately, this thorough vetting process enhances the study’s overall quality and credibility, demonstrating a commitment to creating robust, well-crafted clinical scenarios that closely align with the study’s objectives and can yield valuable research outcomes.

### 2.6 Panel inclusion criteria and recruitment process

Experts from a range of ED settings will be invited to participate in the OxyGuidED study, which focuses on evaluating and rating the oxycodone appropriateness criteria. The panel will comprise experts with diverse expertise in emergency medicine and nurse practitioners who are authorised to prescribe nationally, serving as voting members (*Queensland Health* 2025). Pharmacists will also be encouraged to join the panel as voting members, even though they are not permitted to initiate opioid prescriptions within a national context. Pharmacists with experience in high-risk medication review, analgesia protocols, or opioid safety will be prioritised. The inclusion of at least one pharmacist will bring a system-level perspective that physicians or nurses may overlook, especially in areas such as medication safety, the potential for unsafe interactions among special populations, dispensing logistics, and post-ED continuity of oxycodone treatment. This diverse composition of experts (panellists) aims to ensure that the developed criteria are applicable, reliable, transparent, and tailored. Furthermore, the diverse expertise of the panellists can help identify potential challenges or limitations in implementing the criteria, leading to more refined and adaptable guidelines that can be effectively integrated into various ED workflows and protocols.

The invitation to participate will be sent via email. It will include a Participant Information Form outlining details of the RAND/UCLA Appropriateness Method panel, along with an Expression of Interest form. Prospective panellists will complete an Expression of Interest form through a secure link to the REDCap platform, hosted by QUT (Harris et al., 2009). During the expression of interest phase, applicants are assessed based on their years of clinical experience, including specific involvement with oxycodone, such as prescribing, administering, and reviewing. Additionally, consideration is given to participation in opioid-related committees, relevant education, research activities, and professional memberships in pain societies, emergency medicine, or pharmacy regulatory bodies.

Eligible panellists will then be provided, via a secure REDCap link, with a Consent Form and a Recognition Form to acknowledge future authorship and publication contributions (see the Consent Forms document). Since participation is entirely voluntary, a Withdrawal Form will also be available for panellists who decide to discontinue their involvement at any stage. Contact details of the research team will be provided for any questions or requests for further information. Once consent forms are submitted, the distribution phase of the OxyGuidED survey will commence. Instructions for accessing this platform will be given to all panellists. The evidence review and definitions of key terms will be available for download via REDCap. Links to clinical guidelines and the RAND/UCLA Appropriateness Method rating approach manual will be provided if further information is needed.

## 3. Data Collection, Analysis, and Interpretation

### 3.1. Rating process: First round (Online individual rating)

The rating process will follow the RAND/UCLA Appropriateness Method data collection process and be conducted in two rounds. In the first round, each panellist will independently review a set of scenarios designed to reflect common presentations in adult ED where oxycodone might be considered. Panellists will assess the appropriateness of oxycodone prescription for each scenario using a 9-point Likert scale, where 1-3 indicates inappropriate use (harm outweighs benefits), 4-6 indicates uncertainty (equivocal or unclear benefit-risk balance), and 7-9 indicates appropriate use (benefits outweigh harms) (Fitch et al., 2001). These ratings will be submitted individually via REDCap and can be viewed only by the research team.

Initial data collected from the first round will be analysed to determine the median and the disagreement index for each scenario (Fitch et al., 2001). A summary table and graph will be prepared for each scenario to illustrate disagreements and serve as a reference before the second-round discussion.

### 3.2. Rating process: Second round (Face-to-face discussion and rating)

In the second round, panellists will engage in a moderated in-person discussion to review the anonymised, aggregated distribution of first-round ratings, discuss scenarios with significant disagreement or uncertainty, and consider additional clinical insights or evidence. The moderators will inform the experts before the second round that the discussion should be conducted with respect and that reaching consensus is not obligatory. Furthermore, the moderators will motivate the panellists to engage actively in the discussion and ensure the participation of all panellists. The second round will be overseen by a non-voting member with experience in using the method.

During the second-round discussion, scenarios may be clarified, amended, or merged based on panellists’ feedback and observed patterns of disagreement (Fitch et al., 2001). Where appropriate, new scenarios may also be introduced to address gaps identified during the deliberation process. These refinements ensure recommendations reflect nuanced clinical judgment. This structured approach does not aim to achieve consensus among experts; rather, it facilitates comprehensive discussion and consideration of diverse clinical perspectives. This process ultimately leads to more informed and reliable recommendations for the use of oxycodone in adult ED settings.

Following this discussion, panellists will re-rate each scenario using the same 9-point scale. The rating scores will be recorded on a digitally secured platform (Excel) that will be shared between panellists and the research team.

### 3.3. Data management and analysis

Data received will be stored securely on a server protected by a password, managed by the principal investigator. When co-investigators require the sharing of information, datasets will be exchanged via workplace email and secure digital platforms, utilising password protection. Any printed copies of study documents will be secured in a locked filing cabinet within the primary investigator’s secured office. Data will be anonymised, with only their personal and then aggregated ratings available to the panellists.

The ratings obtained from the second round of the expert panel will be analysed using standard RAND/UCLA Appropriateness Method procedures (Fitch et al., 2001). The final appropriateness of each scenario will be determined based on the median score and the presence or absence of disagreement. For each clinical scenario, the median score will be calculated to represent the central tendency of panellists’ judgments. In addition, disagreement will be assessed using the interpercentile range (IPR) and the interpercentile range adjusted for symmetry (IPRAS), which account for both the spread and symmetry of ratings across the panel. The IPR represents the range between the 30th and 70th percentiles of scores, while the IPRAS adjusts this range based on the degree of symmetry in the distribution around the median. Disagreement is considered present when the IPR exceeds the calculated IPRAS for a given item (Fitch et al., 2001).

Based on the combination of median score and presence or absence of disagreement, each scenario will be classified as:

- Appropriate: Median score of 7–9, without disagreement
- Uncertain: Median score of 4–6 or any median with disagreement
- Inappropriate: Median score of 1–3, without disagreement

This structured approach enables a nuanced and statistically informed interpretation of expert panel ratings, striking a balance between consensus and divergence in clinical judgment.

### 3.4. Interpretive synthesis and evidence mapping

Following the expert panel’s rating and ranking of the criteria, an interpretive synthesis will be conducted to integrate quantitative data from the RAND/UCLA Appropriateness ratings with qualitative insights from the panel discussion and supporting literature. This synthesis aims to identify patterns of agreement or disagreement among panel members and to interpret how related factors affect oxycodone prescribing decisions in the ED.

The evidence map will then be created to visualise the connection between appropriateness ratings, supporting evidence levels, and the clinical domains identified in the review. This mapping will provide a clear overview of how evidence and expert judgment converge to form the final OxyGuidED framework, ensuring that recommendations are grounded in both empirical data and an understanding of the emergency practice context.

## 4. Methodological Rigour and Data Integrity

This study adheres to a detailed protocol that aligns with the RAND/UCLA Appropriateness Method (Fitch et al., 2001), which will be followed to ensure transparency, reproducibility, and minimise bias. The generated scenario will undergo expert review and pilot testing to confirm its clinical realism, clarity, and content validity. Panel members will be selected based on predefined criteria to ensure appropriate expertise and representation, and the generated OxyGuidED criteria are expected to achieve high accuracy. The OxyGuidED survey will be sent to panellists via a personalised link to prevent record duplication, ensuring each panellist has only one entry. Data will be collected using the REDCap platform, which features a mandatory field requirement to avoid missing or invalid entries. The research team will independently verify exported data after each round for accuracy before conducting analysis. Data will be analysed using the RAND UCLA Appropriateness method analysis guidelines provided by Fitch et al. (2001), and it will be published solely based on the aggregated appropriateness rating.

## 5. Discussion

This study aims to develop evidence-based, expert-endorsed criteria for the appropriate prescription of oxycodone in adult ED settings. By employing the RAND/UCLA Appropriateness Method, the findings will assist clinicians in making more consistent, safe, and judicious decisions regarding oxycodone prescriptions in acute pain in the ED setting. The OxyGuidED criteria can provide stakeholders with new insights and inform the revision of opioid prescription guidelines. The objective of developing these criteria is not merely to decrease the prescription of oxycodone but to establish a balance that achieves treatment objectives without inflicting harm, particularly in the ED settings, which have experienced issues of either opioid misuse or undertreatment of pain. These criteria can serve as a robust foundation for institutional guidelines, government recommendations, clinical decision support tools, and educational or training interventions aimed at reducing unnecessary opioid exposure while ensuring adequate pain management. Achieving this balance requires revising opioid prescribing guidelines to align with the Australian National Stewardship Program (*Australian Commission on Safety and Quality in Health Care*, 2022).

The methodology and framework established in this protocol paper can also be adapted to evaluate the overuse of other high-risk medications or unnecessary procedures, thereby supporting ongoing quality improvement and patient safety efforts across all healthcare settings. However, this initiative will establish a robust foundation for future research into alternatives to inappropriate or uncertain oxycodone prescription practices. Future research could investigate the applicability, acceptability, and effectiveness of the developed criteria across diverse health contexts.

## 6. Strengths and Limitations

The RAND/UCLA Appropriateness Method is a notable strength of this study, as it systematically integrates published evidence with expert clinical judgment in areas where extensive empirical data are lacking. Its structured rating process, incorporation of evidence summaries, and controlled feedback across multiple rounds enhance transparency, enabling the documentation of both consensus and disagreement without enforcing consensus. This approach develops context-specific, practice-relevant criteria that reflect real-world complexities while maintaining confidentiality. Owing to its transparency, reproducibility, and practicality, this method holds significant value for ED shared prescribing decisions, particularly for high-risk medications such as oxycodone.

A key limitation of the RAND/UCLA Appropriateness Method is its reliance on structured expert judgment, which, while systematic, remains inherently subjective. Results will reflect the perspectives of clinicians practicing in Australian EDs, which may limit transferability to other settings. The validity of the resulting criteria depends on the panel’s expertise and diversity, and the purposive selection of panellists may raise concerns about selection bias in other methodological contexts. However, these limitations can be mitigated through careful panel composition, transparent documentation of the method, and the use of clear, well-developed clinical scenarios. The iterative design of the RAND/UCLA Appropriateness Method, particularly its structured discussion phase, fosters critical reflection and reduces the risk of unchecked individual bias. These constraints are inherent to the method rather than the study execution and will be acknowledged when interpreting the criteria.

## 7. Conclusion

The methodology outlined in this protocol provides a rigorous and transparent framework for developing criteria to guide the appropriate use of oxycodone in adult ED settings. Once established, these criteria have the potential to support evidence-informed decision-making, improve the quality and consistency of appropriate oxycodone prescribing, and reduce inappropriate use. The resulting guidance may serve as a valuable clinical resource applicable across diverse practice settings, both nationally and internationally.

## Supporting information

(See the Letter document)

(see the Consent Forms document)

## Data Availability

No dataset were generated or analysed for this study protocol. Data will be generated during the subsequent phases of the study and will be reported in future publication.

## 8. Declaration

### 8.1. Ethics approval

Ethical approval for this study has been secured from Queensland University of Technology, with the ethical approval reference (LR 2025-9882-25374). This study is classified as low risk and does not compromise the confidentiality or privacy of data provided by the panellists (See the Letter document).

### 8.2. Consent for publication

Not applicable.

### 8.3. Availability of data and materials

No data have yet been collected for this study.

### 8.6. Author’s contributions

All authors participated in the formulation and design of the study, contributed to the development of the research protocol, and were involved in preparing the manuscript. All authors have reviewed and approved the final version of this manuscript.

## 8.7. Acknowledgment

The author expresses gratitude to the SACM for their financial assistance, as well as to QUT and the supervisors for their guidance and academic support.

