## Supplementary material for "Developing Evidence-Based Criteria for Oxycodone Prescribing in Emergency Departments: A Protocol for the OxyGuidED Study Using the RAND/UCLA Appropriateness Method": (See the Letter document)

1 August 2025

Dear Mrs Suhair Mohsen M Althagafi,

We are pleased to advise that your application has been reviewed and approved by the University Human Research Ethics Committee (UHREC) or delegated review body as meeting the requirements of the National Statement on Ethical Conduct in Human Research (2023).

| | |
|---|---|
| **Project title:** | Development of an evidence-based criteria for appropriateness of oxycodone use in adult ED for treatment of pain (OxyGuidED Study) |
| **Approval number:** | 9882 |
| **Approved version:** | LR 2025-9882-25374 |
| **Approval date:** | |
| **Expiry date:** | |

**Documents approved:**

| Document Type | File Name | Date | Version |
|---|---|---|---|
| Default | Invitation email version 2 | 24/07/2025 | 02 |
| Default | Participation information form-submission | 24/07/2025 | 02 |
| Default | Interest to Participate Form | 24/07/2025 | 01 |
| Default | protocol-Ethics-submission | 24/07/2025 | 02 |
| Default | reviewer comments | 20/07/2025 | 01 |
| Default | Invitation email. tracked changes | 25/07/2025 | 02 |
| Default | Participation information form-track changes | 25/07/2025 | 02 |
| Default | Protocol track changes | 25/07/2025 | 02 |

**Research team approved:**

Dr James Hughes

MRS Suhair Mohsen M Althagafi, Prof Samantha Keogh

This approval is subject to the standard conditions of approval as well as any additional conditions of approval indicated by the UHREC or delegated review body.

**Additional conditions of approval:**

- Nil

Kind regards,

Office of Research Ethics and Integrity

**QUT Human Research Ethics Advisory Team** | | +61 (0)7 3138 5123

** This is a system generated email - PLEASE DO NOT REPLY TO THIS MESSAGE **
