## Supplementary material for "Developing Evidence-Based Criteria for Oxycodone Prescribing in Emergency Departments: A Protocol for the OxyGuidED Study Using the RAND/UCLA Appropriateness Method": (see the Consent Forms document)

| 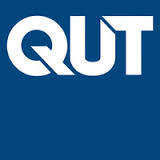 | **CONSENT FORM FOR QUT RESEARCH PROJECT** |
| --- | --- |
| Development of an evidence-based criteria for appropriateness of oxycodone use in adult ED for treatment of pain (OxyGuidED Study)  QUT Ethics Approval Number (LR 2025-9882-25374) | |

**Research team:**

| **Primary Investigator** | **Suhair Althagafi** | **+61402089762** | |
| --- | --- | --- | --- |
| **Principal Investigator** | **James Hughes** | **07 31383830** | |
| **Associate investigator** | **Samantha Keogh** | **3138 3881** | |

**Statement of consent**

**By signing below, you are indicating that you:**

- Have read and understood the information document regarding this research project.
- Have had any questions answered to your satisfaction.
- Understand that if you have any additional questions, you can contact the research team.
- Understand that you are free to withdraw without comment or penalty.
- Understand that if you have concerns about the ethical conduct of the research project you can contact the Research Ethics Advisory Team on +61 7 3138 5123 or.
- Understand that the research project will include an audio and/or video recording.
- Agree to participate in the research project.

**Please tick the relevant box below:**

**☐ I agree for the panel meeting to be audio / video recorded.**

**☐ I do not agree for the panel meeting to be audio / video recorded.**

| Name |
| --- |
| Signature |
| Date |

| 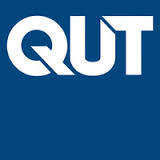 | **CONSENT FORM FOR Panellist Recognition in Publication** |
| --- | --- |
| Development of an evidence-based criteria for appropriateness of oxycodone use in adult ED for treatment of pain (OxyGuidED Study) | |

Thank you for your valuable contribution to our research project. We would like to acknowledge your efforts in our upcoming publication. Please indicate your preference for recognition by completing this form.

**Participant Information:**

- **Name: ________________________________________________________**
- **Affiliations:**
- **________________________________________________________________**
- **________________________________________________________________**
- **________________________________________________________________**
- **Email: _________________________________________________________**

**Consent Options: Please select one of the following options:**

**☐** I wish to be listed as a co-author in the publication**.**

**☐** I wish to be acknowledged in the publication.

**Statement of Consent:**

I, ________________________________, hereby give my consent to be recognized as indicated above in the publication titled "Development of an evidence-based criteria for appropriateness of oxycodone use in adult ED for treatment of pain (OxyGuidED Study)". I understand that this recognition is based on my contribution to the research project.

**Signature: __________________________
Date: _______________________________**

**The following form is OPTIONAL**

| 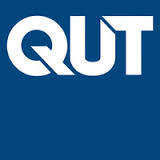 | **WITHDRAWAL OF CONSENT FOR QUT RESEARCH PROJECT** |
| --- | --- |
| Development of an evidence-based criteria for appropriateness of oxycodone use in adult ED for treatment of pain (OxyGuidED Study)  **QUT Ethics Approval Number (**LR 2025-9882-25374) | |

**Research team**

| **Primary Investigator** | **Suhair Althagafi** | **+61402089762** | |
| --- | --- | --- | --- |
| **Principal Investigator** | **James Hughes** | **07 31383830** | |
| **Associate investigator** | **Samantha Keogh** | **3138 3881** | |

I hereby wish to WITHDRAW my consent to participate in the research project named above.

I understand that this withdrawal WILL NOT jeopardise my relationship with QUT.

**☐ I request that data collected about me that is still identifiable is not used in data analysis.**

**☐ Please use data collected about me so far for the study.**

| Name |
| --- |
| Optional: Signature |
| Date signed or noted |
| Optional:  Reason for withdrawal |

**Please note that you may also withdraw verbally or via email.**
